# Introduction of a European Central-South-Eastern West Nile Virus Lineage 2 strain in Italy in 2023: evidence from the first locally acquired neuroinvasive case in the Calabria region

**DOI:** 10.64898/2025.12.19.25342062

**Authors:** Simone Malago, Antonio Mori, Michela Deiana, Maria Vittoria Mauro, Valeria Vangeli, Giuliana Guadagnino, Silvia Accordini, Natasha Gianesini, Lorena Maria Chesini, Samuele Cheri, Sonia Greco, Francesca Greco, Jesse Julian Waggoner, Chiara Piubelli, Federico Giovanni Gobbi, Concetta Castilletti, Antonio Mastroianni

**Author notes:** These authors share correspondence. **Correspondence**: Dr. Chiara Piubelli, Department of Infectious, Tropical Diseases and Microbiology, IRCCS Sacro Cuore Don Calabria Hospital, via Don A. Sempreboni, 5, Negrar di Valpolicella (VR), 37024, Italy., Dr. Concetta Castilletti, Department of Infectious, Tropical Diseases and Microbiology, IRCCS Sacro Cuore Don Calabria Hospital, via Don A. Sempreboni, 5, Negrar di Valpolicella (VR), 37024, Italy. These authors contributed equally to this work and share last authorship (Concetta Castilletti, Antonio Mastroianni).

## Abstract

**Objective:** West Nile virus lineage 2 (WNV-2) is a growing public health concern in Europe causing West Nile fever or West Nile neuroinvasive disease (WNND) with substantial morbidity and mortality; however, genomic data from Southern Italy are limited despite recent expansion of autochthonous transmission. The aim of the study was to characterise the phylogenetic and molecular features of the WNV-2 strain responsible for the first autochthonous human infection reported in Calabria (2023), and two more additional WNND cases detected in 2024.

**Methods:** Full WNV-2 genomes were generated from the three cases. Phylogenetic analysis was performed using all publicly available WNV sequences up to September 2025. Amino-acid changes in the polyprotein were compared with known WNV-2 lineage and sub-lineage signatures.

**Results:** The three sequences formed a monophyletic group within sub-lineage WNV-2a, clustering with strains circulating in Central-South-Eastern Europe and showing closest affinity to Hungarian sequences. Non-synonymous substitutions characteristic of the Hungary 578/10 ancestor (NS2B-119I, NS4B-14G, NS4B-49A and NS5-298A), were identified and absent from Central-Northern-Western European and previously reported Italian sequences. Additional substitutions (E-159T, E-399R and NS3-249P) corresponded to signatures from a fatal WNV-2 infection in a Great Grey owl in Slovakia.

**Conclusions:** Our study provides the first report of Central-South-Eastern European WNV-2 circulation outside Eastern Europe supporting its likely spread through the Balkans into Italy by 2022. These findings underscore the rapid spread of WNV-2 in newly affected areas and highlight the critical need for sustained molecular surveillance.

## Introduction

West Nile virus (WNV; species *Orthoflavivirus nilense*) is a mosquito-borne orthoflavivirus, mainly transmitted by *Culex spp*. mosquitoes. Humans are incidental dead-end hosts, and an estimated 75-80% of WNV human infections are asymptomatic, hampering accurate assessment of WNV circulation. Among symptomatic patients, most experience a self-limiting febrile illness termed West Nile Fever. Approximately 1% of infections progress to West Nile neuroinvasive disease (WNND), manifesting as meningitis, encephalitis or acute flaccid paralysis [1,2].

WNV circulation in Europe continues to increase, progressively expanding into previously non-endemic areas both in northern and southern regions (**Figure 1 A-D**, [3]) [4–6]. The emergence of cases in new areas highlights ongoing geographic expansion, likely driven by environmental, climatic and ecological changes [7,8]. Italy is one of the most affected European countries, with a significant proportion of locally acquired infections, WNND cases, and fatalities. The WNND case fatality rate remains high, at 14-20% since 2018. In 2025, WNV was detected across 53 provinces in 14 regions, with Lazio and Campania reporting a significant increase in cases (252 and 124 cases, respectively, **Figure 1 E-H**, [9]) [9].

**Figure 1.**
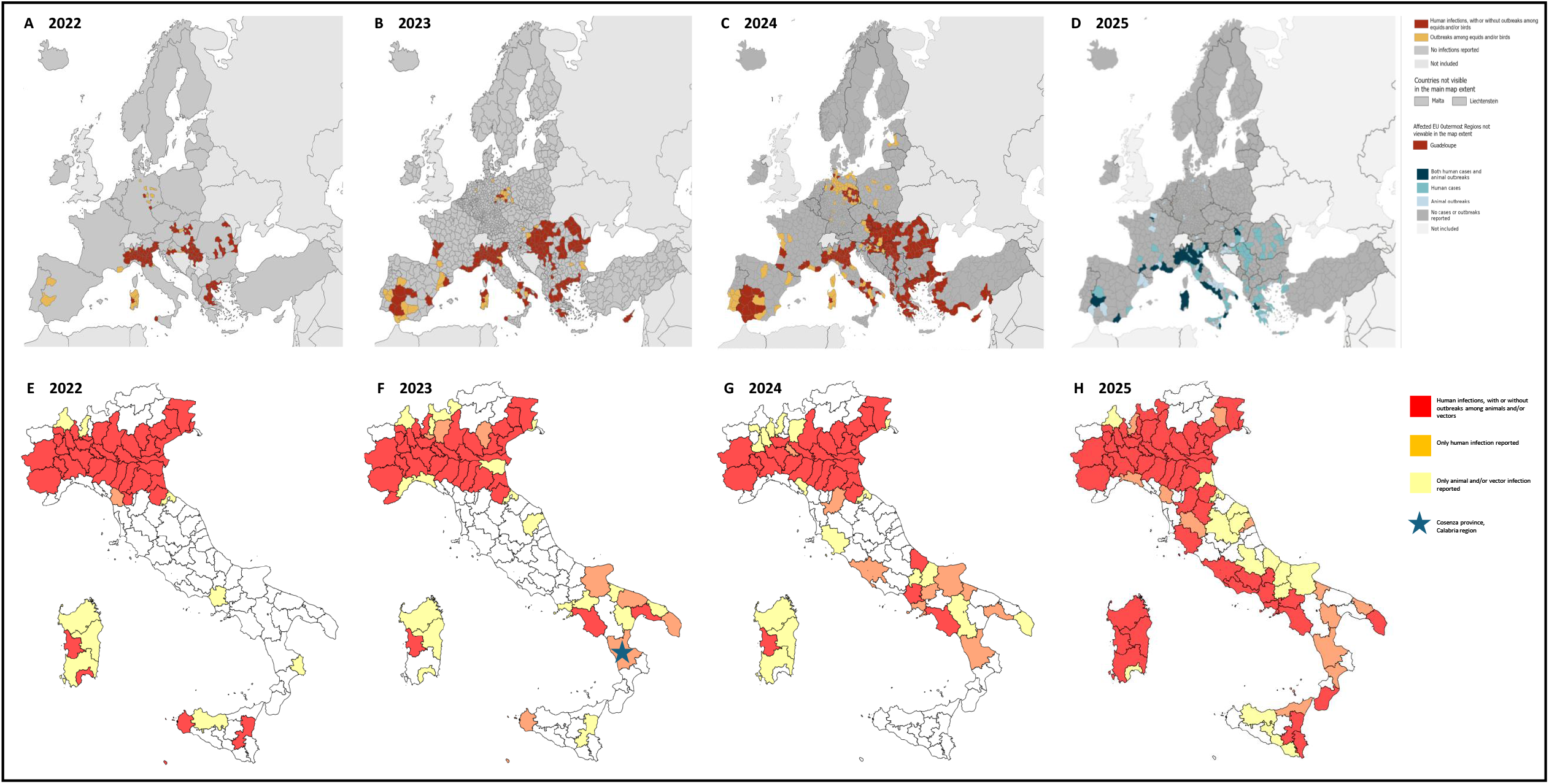
Spatial distribution of animal and human WNV infections in the EU/EEA and neighboring countries from 2022 to 2025. Spatial distribution of animal and human WNV infections in the EU/EEA and neighboring countries from 2022 to 2025: A) 2022; B) 2023; C) 2024; D) 2025 as of 8 October; and spatial distribution of animal, vector and human WNV infections in Italy: E) 2022; F) 2023; G) 2024; H) 2025 as of 29 October 2025. Adapted from European Centre for Disease Prevention and Control, European Food Safety Authority (ECDC/EFSA database) [3] and from Istituto Superiore di Sanità [9]. Surveillance of West Nile virus infections in humans in Europe distribution is reported in NUTS 3 (nomenclature of Territorial Units for statistics, provinces level) or GAUL 1 (Global Administrative Unit layers, subnational level 1) regions of the EU/EEA and neighboring countries.

WNV is characterized by high genetic diversity, with at least nine lineages worldwide. Lineages 1 (WNV-1) and 2 (WNV-2) are the most widespread and most frequently associated with human cases [1,10]. WNV-2 currently predominates in Europe [11,12], and two main sub-lineages have been identified: WNV-2a and WNV-2b. The most prevalent in Europe, WNV-2a, can be further divided into two clusters, referred to here as the Central-North-Western (CNW) and Central-South-Eastern (CSE) clusters, respectively. CNW likely emerged in Austria around 2006 and has been detected mainly in Germany and northern Italy, whereas CSE, appears to have originated in Hungary around 2007 with a similar evolution rate, and includes strains circulating in Greece and other Central and South-Eastern European regions [6]. Continuous molecular characterization is essential to elucidate the complex viral dynamics of WNV lineages and clusters, monitor the possible introduction of more virulent/pathogenic strains, and clarify the pathogenetic mechanisms.

In September 2023, a patient presenting with severe neurological symptoms was admitted to the “Annunziata” Hub Hospital, Azienda Ospedaliera di Cosenza, in the Calabria region, the southernmost region of peninsular Italy, where laboratory testing confirmed WNV infection. This represented both the first autochthonous WNV case and the first WNND in Calabria. Here, we describe the molecular and phylogenetic features of the WNV strain responsible for this case as well as two additional cases detected in 2024.

## Materials and Methods

This study included the full genome characterization, phylogenetic and variant analysis of WNV derived from the three described WNND human cases, diagnosed in the Calabria Region (South Italy) in 2023-2024. Details about laboratory diagnosis, next generation sequencing and bioinformatic analysis are reported in dedicated section of Supplementary material.

### Ethical statement

Ethical approval was not required according to the *National Plan for the Prevention, Surveillance, and Response to Arboviruses (PNA) 2020-2025* [13]. Human samples were collected by clinicians and processed by the personnel of the Pathology Unit at the “Annunziata” Hub Hospital. All data were analysed anonymously following the PNA guideline.

## Results

### Genome analysis of WNND cases detected in Calabria (2023–2024)

Blood from the WNND case diagnosed in September 2023 tested positive for WNV RNA, with a viral load of 11,155 copies/mL. The two WNND cases diagnosed at the same hospital in August and September 2024 had urine samples, with viral loads of 842,845 copies/mL and >50,000,000 copies/mL, respectively. Viral genomes were then sequenced, and for the lower viral load blood sample, two complementary approaches were used based on hybrid capture and amplicon-based sequencing.

### Consensus generation

For the 2023 case, reads generated from both sequencing methods were merged and aligned against the sequence of a WNV-2 isolated in the Campania region in November 2024 (GenBank ID: PQ654050). This produced a breadth of coverage of 88% at 5X and 77% at 10X. Sequencing reads from the two 2024 cases showed >99% genome coverage at 10X when aligned to the same reference. The resulting consensus sequences (WNV_IRCCS-SCDC_01/2025_, WNV_IRCCS-SCDC_02/2025_ and WNV_IRCCS-SCDC_03/2025,_ respectively) were used for phylogenetic analysis. Complete sequencing metrics are reported in **Table S1**.

### Phylogenetic analysis

Phylogenetic analysis showed that all three genomes belonged to the WNV-2, sub-lineage 2a, clustering within sequences from the European Central-South-Eastern cluster (**Figure 2, panel A**). Neighboring branches included sequences from Greece, Hungary, Serbia, Russia, Romania, Slovakia, Poland and Kosovo [6], with the most closely related genomes originating mainly from Hungary. Only two of the nearest sequences were associated with countries other than Hungary, both linked to Spain. Among Italian genomes, the closest sequences were from southern-Italy, mainly from the Campania region in 2022-2025. By contrast, all sequences from northern and central Italy clustered on a separate branch together with those from Sardinia, while adjacent branches were mainly composed of strains from Central-North-Western Europe, with additional genomes from other Eastern European countries.

**Figure 2:**
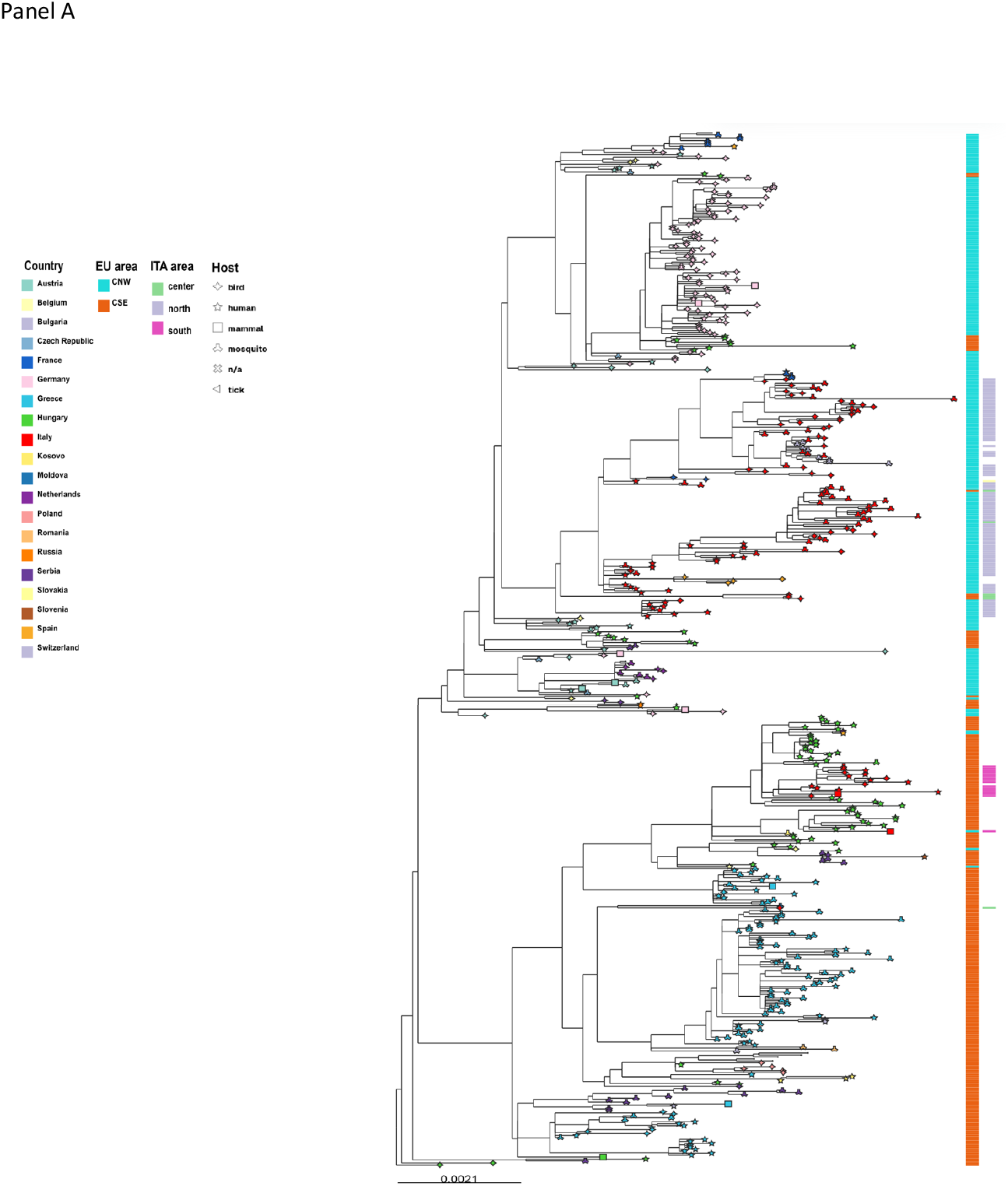
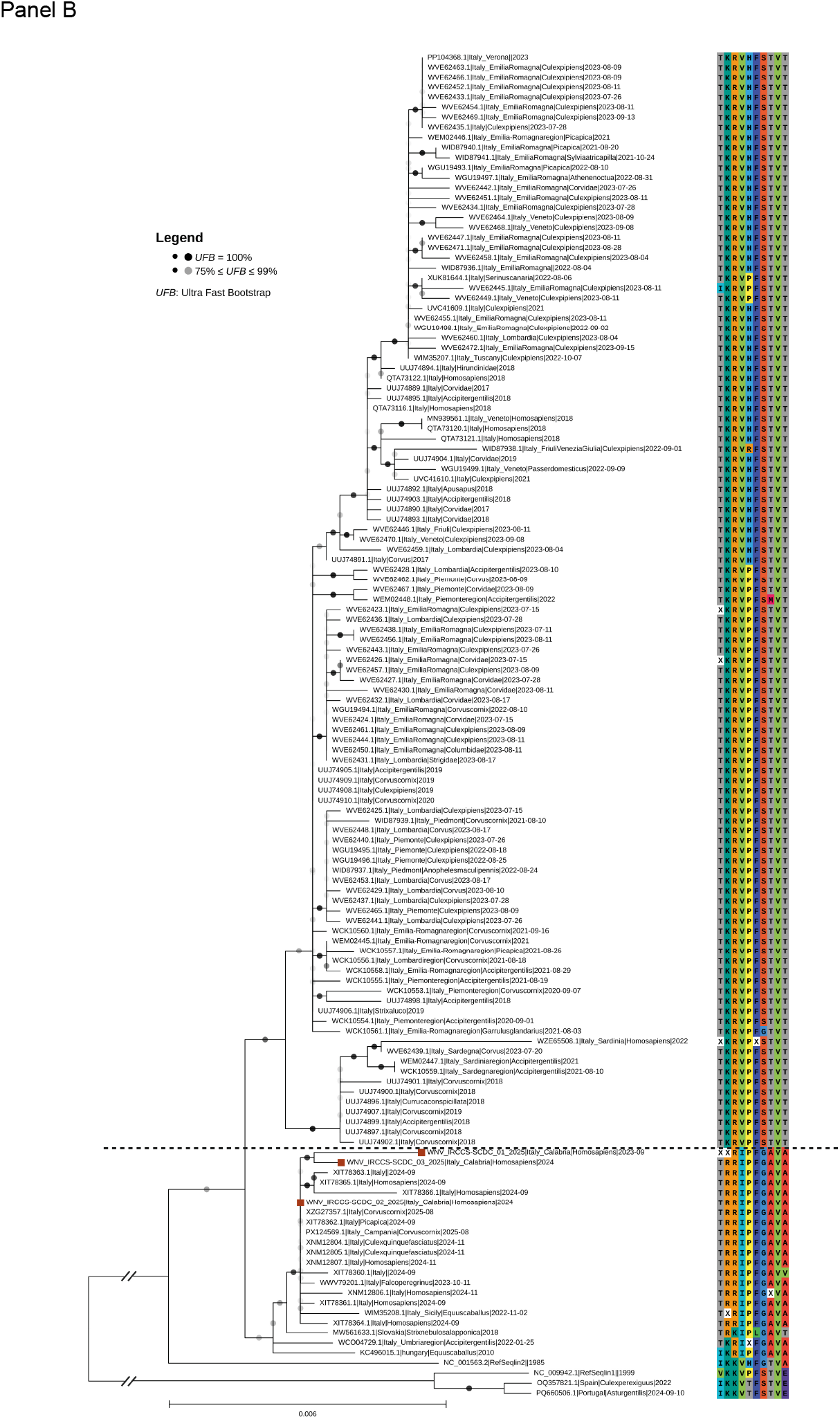
Phylogenetic analysis results. **Panel A: Maximum likelihood phylogenetic tree of the European WNV-2 strains**. Leaves shapes represent the type of host. Colors of the leaves represent the country of collection. Geographical areas refer to the columns on the right, describing European and Italian sub-areas of WNV-2 distribution. Outgroups and other sequences are excluded for visualization purpose. This sub-tree is part of an African-European tree (**supplementary Figure S1**). **Panel B: Phylogenetic tree of viral strains circulating in Italy, based on complete WNV lineage 2 polyprotein sequences**. Three WNV lineage 1 sequences were included as outgroup. The Hungarian KC496015.1 and Lineage 2 reference sequences were added for comparison. On the right, amino acidic residues at positions of interest for each sequence are displayed. More specifically, from left to right are positions (gene): 449 and 689 (E), 835 (NS1), 1493 (NS2B), 1754 and 1991 (NS3), 2287, 2322 and 2386 (NS4B), 2827 (NS5) of the polyprotein. Sequence labels include strain identifiers, country of collection, host and date of sampling. Dots on tree branches represent bootstrap values as per legend. Our three sequences are indicated with red squares on respective leaves.

### Variant analysis

Variant calling and annotation were performed using the WNV lineage 2 reference genome (GenBank ID: NC_001563.2). A total of 99 Single Nucleotide Variants (SNVs) were identified in WNV_IRCCS-SCDC_01/2025_, with 89 annotated as synonymous and 9 as non-synonymous substitutions. Regarding the 2024 sequences, 197 SNVs were detected in WNV_IRCCS-SCDC_02/2025_ (171 synonymous, 19 non-synonymous), and 138 SNVs were identified in WNV_IRCCS-SCDC_03/2025_ (116 synonymous, 15 non-synonymous). No identified variants were predicted to have high impact by Snp-Eff. The three WNV-2 genomes detected in Calabria region were characterized by a set of specific amino acid residues associated with south-Italian strains, clearly separating them from the northern Italian strains (E-399R, NS2B-119I, NS4B-14G, NS4B-49A and NS5-298A, **Figure 2, panel B**). Furthermore, the E-159T and NS3-249P, two residues of particular interest, was shared by all south-Italian cases and by certain percentage of north-Italian ones.

## Discussion

This study investigated the phylogeny and molecular characteristics of WNV strains responsible for the first autochthonous infections reported from the Calabria region, which resulted in WNND. Phylogenetic analysis showed that the three WNV-2 genomes belong to a monophyletic clade within sub-lineage 2a that derived from a single introduction into Europe of the prototype Hungarian strain (Hungary 2004, GenBank Accession ID: DQ116961). These clustered with CSE strains, and the most closely related Italian sequences originated from southern Italy, mainly Campania between 2022 and 2025.

Our report provides the first evidence of the circulation of a Hungary 578/10 WNV-2-like strain outside of Eastern Europe. This strain appears to have spread across Central-South-Eastern Europe over the past five years, reaching Italy through the Balkans likely in 2022, leading to the emergence of a WNV-2 strain with potentially key adaptive mutations. In the Calabrian cases, we identified non-synonymous substitutions representative of Hungary 578/10, originally isolated in 2010. In particular, amino acidic substitutions NS2B-119I, NS4B-14G, NS4B-49A and NS5-298A, were not found in European CNW sequences (including those from North-Central Italy, **Figure 2, panel B**). Additionally, E-159T, E-399R, and NS3-249P were identified as signatures of the southern-Italian cases, in common with a case of WNV neuroinvasive case in a Great Grey owl (Slovakia 2018, GenBank Accession ID: MW561633.1) [14].

The proline at position NS3-249 has been previously associated with increased neurovirulence [15,16]. In comparison to CSE strains, approximately half of the sequences from northern Italy carry a valine at this position (**Figure 2, panel B**). NS3 encodes the helicase protein, and a proline at position 249 enhances mortality in birds and increases viremia, facilitating virus transmission [15]. In silico structural modeling suggests that NS3-249P also uniquely retains activity at 42°C (avian body temperature) likely contributing to greater protein stability and viral transmissibility [15,16].

NS3-249P has independently emerged in WNV-1 and -2 strains and preceded human outbreaks in geographically distant countries over the past 70 years [15,17]. Furthermore, it may contribute to differences in mortality among human WNND cases in the CSE cluster (higher) versus the CNW cluster (lower) [18]. Substitution E-159T in the envelope protein is considered a determining factor of WNV neurovirulence due to its impact on the host response and neuronal cell degeneration [19,20]. This substitution, absent in the Hungarian 578/10 strain (**Figure 2, panel B**) but present in southern Italian strains, had previously been identified in several Italian WNV-2 genomes isolated between 2011 and 2014 in northern Italy [15,16].

In recent years, southern Italy has experienced an abrupt increase in locally acquired WNV-2 WNND cases, leading to the major epidemic observed in the Lazio and Campania regions in 2025 (**Figure 1**) [21–24]. Although preliminary data do not allow us to determine whether the detected Calabria strains exhibit enhanced neurovirulence, further investigations are warranted to clarify this point. Overall, our findings contribute to a better understanding of WNV-2 transmission dynamics in Europe and highlight the need for sustained molecular surveillance to monitor viral evolution and diversity.

## Data availability

Sequences raw reads were uploaded in the European Nucleotide Archive (ENA, EMBL-EBI) (project code PRJEB104938). Consensus sequences for WNV_IRCCS-SCDC_01/2025_, WNV_IRCCS-SCDC_02/2025_ and WNV_IRCCS-SCDC_03/2025_ were uploaded in GenBank (NCBI, accession IDs: PX289511, OZ374057.1, OZ374058.1 respectively)

## Transparency declaration

Authors do not have conflicts of interests to declare for this work.

## Authors’ contributions

SM, AMo, MD, CP, and CC contributed to the conceptualisation of the manuscript, interpreted results and drafted the manuscript. MVM, FG, and performed specific diagnostic tests. SM, AMo, MD, CP, and CC analysed and interpreted data. SG, VV, GG and AMa, were responsible for the clinical management of the patients. CP, FG, FGG, JJW, CC and AMa contributed to the conceptualization and revision of the manuscript. All co-authors contributed to the revision of the manuscript and approved the final version for submission and fulfil all four ICMJE authorship criteria.

