## Supplementary material for "Introduction of a European Central-South-Eastern West Nile Virus Lineage 2 strain in Italy in 2023: evidence from the first locally acquired neuroinvasive case in the Calabria region"

Supporting information alongside the article [**Introduction of a European Central-South-Eastern West Nile Virus Lineage 2 strain in Italy in 2023: evidence from the first locally acquired neuroinvasive case in the Calabria region.**] by Simone Malagò et al., on behalf of the authors, who remain responsible for the accuracy and appropriateness of the content.

### Materials and Methods

#### Clinical samples and laboratory diagnosis

Laboratory diagnosis of WNND was performed at the "Annunziata" Hub Hospital in Cosenza, Italy. For the first autochthonous case identified in Calabria, collected on September 2023, cerebrospinal fluid (CSF) and blood samples were analyzed using the one-step real time RT-PCR WNV ELITe MGB® Kit (ELITechGroup SAS, Puteaux, France), following the manufacturer's instructions. Both samples tested positive for WNV RNA, with viral loads <500 copies/mL in CSF and 11,155 copies/mL in blood. In addition, two additional WNND cases diagnosed in 2024 at the same hospital were also included. For both patients, urine sample were collected in August and September 2024, respectively; with viral loads 842,845 copies/mL and >50,000,000 copies/mL.

### Ethical statement

Ethical approval was not required according to the *National Plan for the Prevention, Surveillance, and Response to Arboviruses (PNA) 2020-2025* [1]. Human samples were collected by clinicians and processed by the personnel of the Pathology Unit at the "Annunziata" Hub Hospital. All data were analysed anonymously following the PNA guideline.

**Viral full genome sequencing analysis**

Sample pre-processing, sequencing and bioinformatic analyses were carried out at IRCCS Sacro Cuore Don Calabria Hospital (Negrar di Valpolicella, Verona, Italy). Analyses were performed on the same samples used for the laboratory diagnosis.

Both whole-blood and urine samples were diluted 1:4 in PBS to improve extraction efficiency and nucleic acids were extracted using EZ1® DSP Virus kit on the Qiagen EZ1 Advanced XL, (Qiagen, Hilden, Germany). RNA quantity and quality were assessed using the Qubit RNA HS assay kit (Invitrogen, Thermo Fisher Scientific, Inc., Waltham, MA, USA) and the High Sensitivity RNA ScreenTape on the 4200 TapeStation System (Agilent Technologies Inc., Santa Clara, CA, USA).

Due to the low viral load of 2023 sample, two sequencing approaches were employed: (i) Illumina RNA Prep kit with enrichment via the Illumina Viral Surveillance Panel (VSP) hybridization capture probes (Illumina, California, USA); and (ii) a 400 bp tiled-amplicons panel as described by Diagne and colleagues [2]. Amplicons were than processed using Illumina DNA Prep kit. Libraries were loaded onto an Illumina P1 flow cell and sequenced in 2x150 mode, on a NextSeq1000 instrument (Illumina).

Reads from both runs were merged and processed via kraken2 v2.1.3 (https://github.com/DerrickWood/kraken2) to eliminate human reads, then trimmed via fastp v0.23.4 (https://github.com/OpenGene/fastp#fastp) and aligned via bwa-mem2 v2.2.1 (https://github.com/bwa-mem2/bwa-mem2) against the GenBank (NCBI, https://www.ncbi.nlm.nih.gov/genbank/) reference sequence NC_001563.2. This alignment yielded a breadth of coverage of 80% at 5X and 70% at 10X. Consensus sequence was retrieved using iVar consensus v1.4.4 (https://github.com/andersen-lab/ivar) with the following setting parameters "-q 10 –t 0.7 -m 5". Alignment statistics were evaluated through samtools "coverage" and "flagstat" v1.22. Inspection of the alignment with Integrative Genomics Viewer (IGV) v2.16.2 (https://github.com/igvteam/igv) revealed a high number of mismatches, prompting realignment against a more closely related sequence (GenBank accession ID: PQ654050), corresponding to a WNV-2 strain detected in Campania (Italy) in November 2024 (see phylogenetic analyses paragraph). This second alignment produced improved coverage (88% at 5X and 77% at 10X); the corresponding consensus sequence was therefore used for phylogenetic analyses.

For the two 2024 samples, which showed high viral loads, only the Illumina hybrid capture protocol was applied. Consensus sequences were generated following the bioinformatic workflow applied to the 2023 sample.

**Phylogenetic analysis**

All available European and African WNV-2 whole-genome sequences were downloaded from NCBI Virus database (https://www.ncbi.nlm.nih.gov/labs/virus/vssi/#/, last accessed on 05 September 2025; sequence length ≥10.5Kb; nucleotide completeness: complete; ambiguous characters ≤10). The dataset included our newly generated consensus genomes, the downloaded sequences and the WNV-2 reference genome (NC_001563.2). Phylogenetic analysis included WNV sequences deposited in the NCBI Virus database from the earliest European and African sequences through September 2025. Most of these were from Italy (n=124), Greece (n=112), Germany (n=99), Russia (n=87) and Hungary (n=76), and were predominantly associated with mosquito (~36%), human (~29%) and bird (~27%) hosts. Multiple sequence alignment was performed using mafft v7.505 (https://github.com/GSLBiotech/mafft). The resulting Multiple Sequence Alignment (MSA) was used to infer a phylogenetic tree with iq-tree v2.3.5

(https://github.com/iqtree/iqtree2) using WNV lineage 1 sequences as outgroup and applying the following parameters "-B 1000 -alrt 1000 -m MFP". The best-fit model was: GTR+F+I+R4. Based on the resulting tree, the genome showing the closest genetic similarity to our sequences was selected and used as a "new reference" to recover the maximum amount of coverage. The sequence "PQ654050" (GenBank accession ID) was therefore used as reference for final alignment and consensus generation, following the same analytical pipeline described above. Trees were visualized via microreact (https://microreact.org/; https://microreact.org/project/wnv2-first-calabria-wnnd).

**Variant analysis**

Variant calling was performed via bcftools v 1.18-9 (https://github.com/samtools/bcftools) and variants were annotated with SnpEff v5.2c (https://pcingola.github.io/SnpEff/) using the the reference genome NC_001563.2 for annotation purpose. Moreover, amino acidic residue comparison was carried out using the Hungarian strain 578/10 (GenBank accession ID: KC496015) as a reference and representative ancestor of the CSE European WNV-2 cluster.

To carry out this analysis, all available Italian WNV lineage 2 complete polyprotein sequences (length aa>3,400) were downloaded from NCBI Virus database (last accessed on 05 September 2025) and combined with the three sequences generated in our centre. Additional sequences of epidemiological interest were included: the isolate from a Great Grey owl from Slovakia (GenBank accession ID: MW561633) [3], the WNV lineage 2 reference (GenBank accession ID: NC_001563.2) and three WNV lineage 1 sequences used as outgroup. In total, 134 amino acid sequences were included and aligned via mafft v7.505. Tree was visualized via TreeViewer v2.2.0 (https://github.com/arklumpus/TreeViewer) [4].

**Sequencing quality**

As shown in Table S1, sample $WNV_{IRCCS\text{-}SCDC\_01/2025}$ produced approximately 6.3 million reads via the hybrid capture sequencing method (Illumina VSP panel), in 2x150 configuration. The short amplicon panel generated approximately 19 million reads, with amplicons of approximately 400bp in length. Sample $WNV_{IRCCS\text{-}SCDC\_02/2025}$ produced a total of 4,626,870 reads via the hybrid capture sequencing method (Illumina VSP panel), in 2x150 configuration. Sample $WNV_{IRCCS\text{-}SCDC\_03/2025}$ produced a total of 11,810,012 reads via the hybrid capture sequencing method (Illumina VSP panel), in 2x150 configuration.

### Supplementary Tables

**Table S1. Sequencing alignment statistics of the three samples analysed at IRCCS Sacro Cuore Don Calabria.** Statistics refer to the alignment against the sequence with GenBank accession ID: PQ654050.

| Sample | Enrichment method | Mapping reads | Breath of coverage (%1X) | Breath of coverage (%5X) | Breath of coverage (%10X) | Mean depth of coverage (X) |
|---|---|---|---|---|---|---|
| **$WNV_{IRCCS\text{-}SCDC\_01/2025}$** | VSP panel | 394 | 82 | 44 | 13 | 5 |
| | Amplicons | 2,193,316 | 74 | 67 | 61 | 1,300 |
| **$WNV_{IRCCS\text{-}SCDC\_02/2025}$** | VSP panel | 75,555 | 100 | 100 | 99.95 | 99.78 |
| **$WNV_{IRCCS\text{-}SCDC\_03/2025}$** | VSP panel | 18,919 | 99.95 | 99.06 | 98.58 | 97.11 |

**Table S2. Positions of investigated residues of interest in WNV-2 polyprotein.** Rows reporting amino acid residues refer to: the Central-South-Eastern clade ancestor from Hungary (GenBank Accession ID: KC496015.1), a Great Grey owl case of WNND (GenBank Accession ID: MW566133.1) from Slovakia, a representative set of the southern-Italian cases and finally a representative set of the central-northern Italian WNV-2 strains.

| **Gene name** | **E** | | **NS1** | **NS2B** | **NS3** | | **NS4B** | | | **NS5** |
|---|---|---|---|---|---|---|---|---|---|---|
| **Position in gene** | **159** | **399** | **44** | **119** | **249** | **486** | **14** | **49** | **113** | **298** |
| **Position in polyprotein** | **449** | **689** | **835** | **1493** | **1754** | **1991** | **2287** | **2322** | **2386** | **2827** |
| **KC496015.1 (Hungary 578/10)** | I | K | R | I | P | F | G | A | V | A |
| **MW561633.1 (Great Grey Owl, Slovakia 2018)** | T | R | K | I | P | L | G | A | V | T |
| **South-Italy** | T | R | R | I | P | F | G | A | V | A |
| **North-Center Italy** | T | K | R | V | P/H | F | S | T | V | T |

**Table S3: Link to the repository of obtained sequences information**

https://docs.google.com/spreadsheets/d/1CX75kl8m-PaHgS-rl5hyX9m36iTxqCTPvNTaUihvZCA/edit?usp=sharing

**Supplementary Figure**

**Figure S1. Complete phylogenetic tree of African-European whole-genomes of WNV-2 (Panel A) and geographical map representing WNV-2a clusters distribution reported in EU countries (Panel B)**

**A**

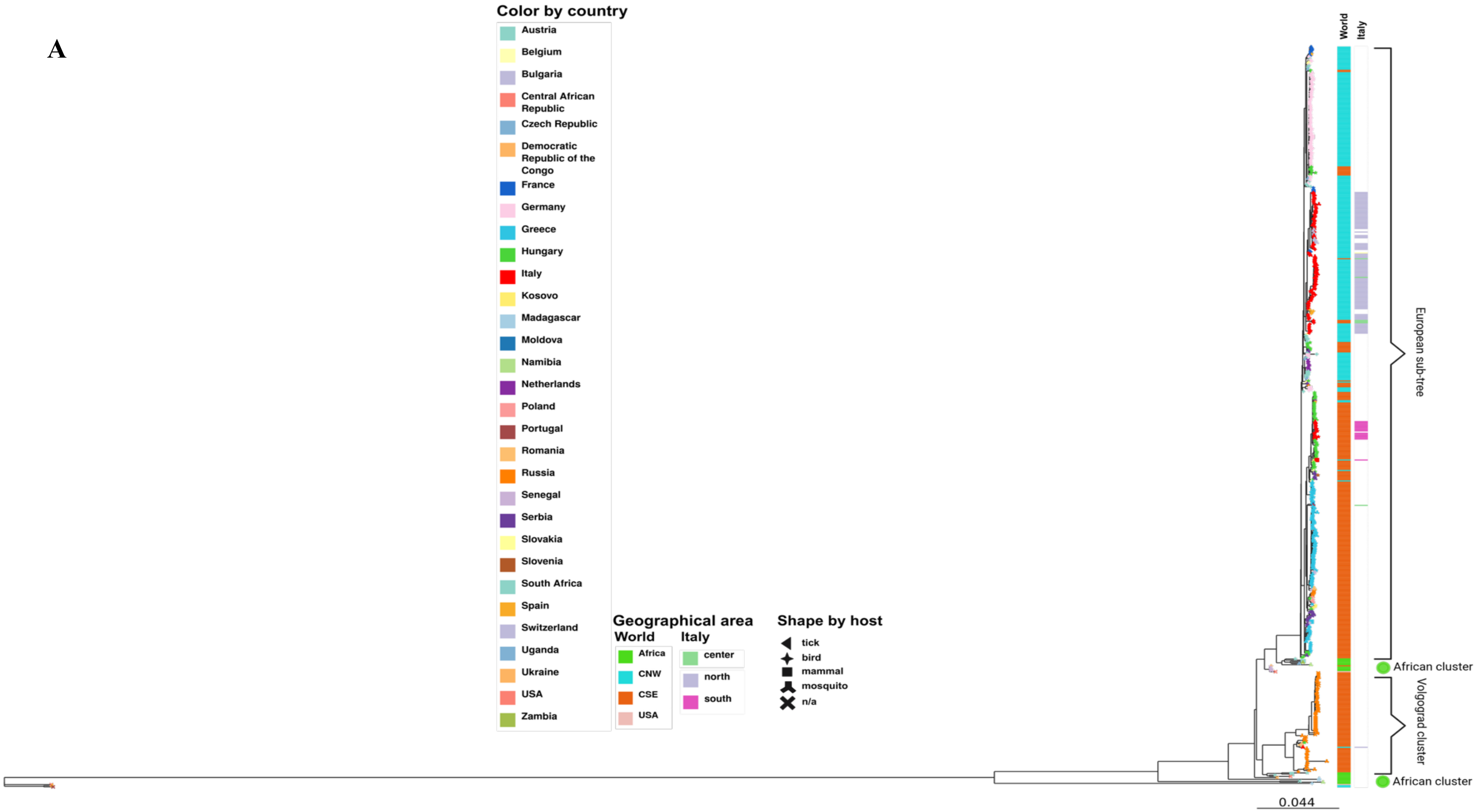

B

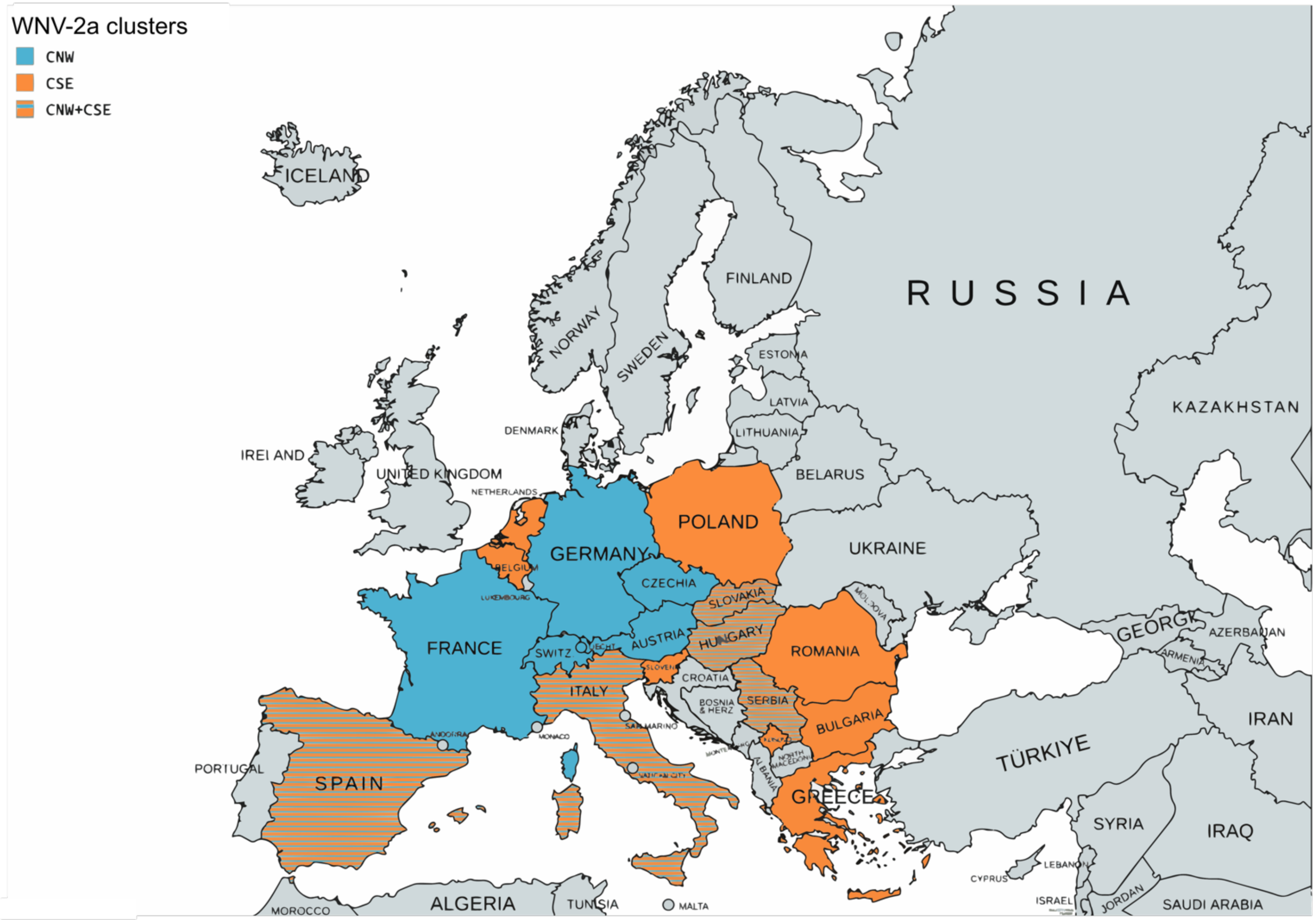


Made with mapchart (https://www.mapchart.net/index.html)

**Figure S2. Phylogenetic sub-tree of the most closely related sequence to the 2023 and 2024 Calabria region's cases.**

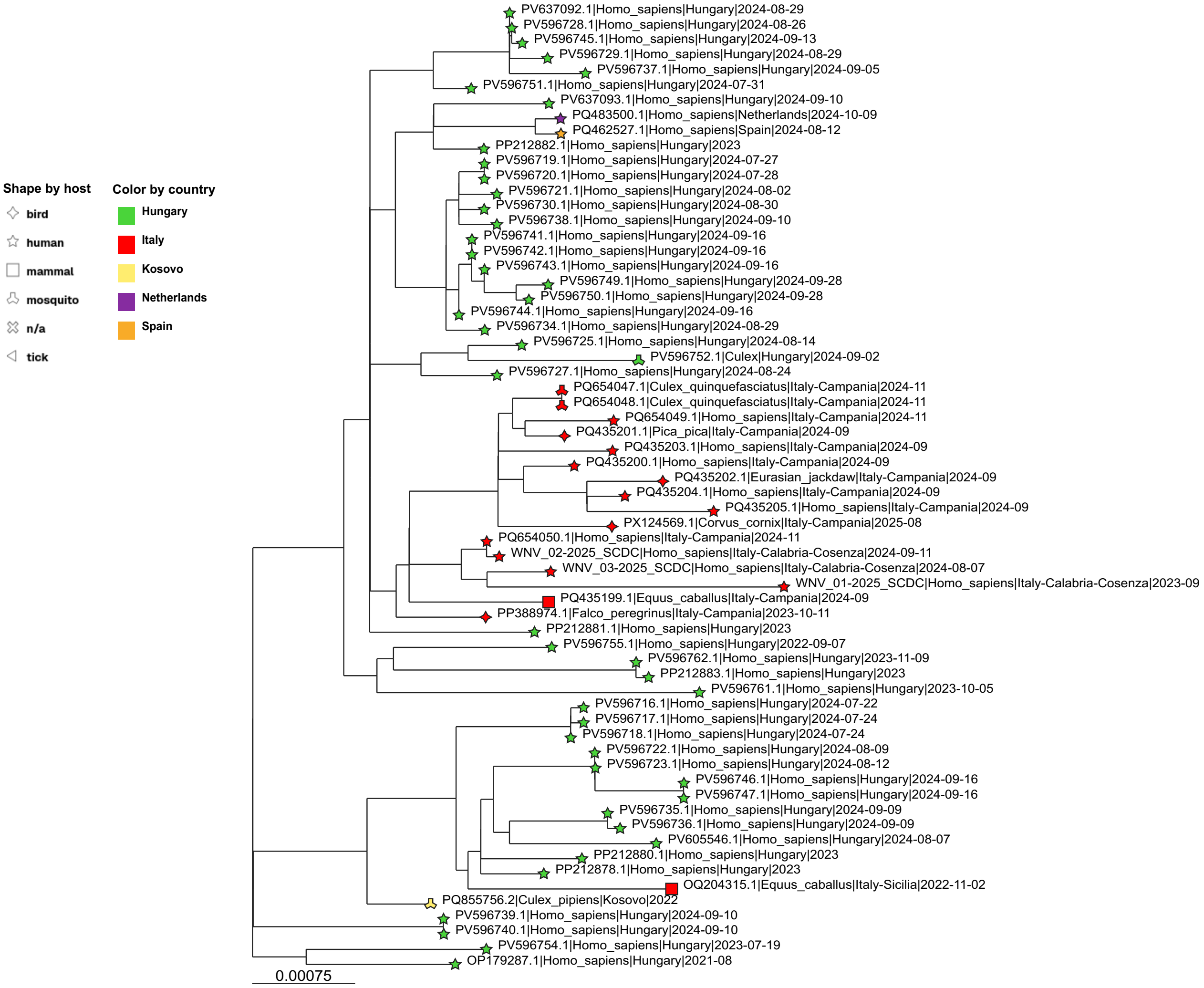


[https://microreact.org/project/wnv2-first-calabria-wnnd](https://microreact.org/project/wnv2-first-calabria-wnnd) (Closest_to_ours)
